# Major depression is not an inflammatory disorder: depletion of the compensatory immunoregulatory system is a hallmark of a mild depression phenotype

**DOI:** 10.1101/2023.12.14.23299942

**Authors:** Michael Maes, Asara Vasupanrajit, Ketsupar Jirakran, Bo Zhou, Chavit Tunvirachaisakul, Abbas F. Almulla

**Author notes:** **Corresponding author:** Prof. Dr. Michael Maes, M.D., Ph.D., and Prof. Dr. B. Zhou, M.D., Ph.D. Sichuan Provincial Center for Mental Health, Sichuan Provincial People’s Hospital, School of Medicine, University of Electronic Science and Technology of China Chengdu 610072, China and Department of Psychiatry, Faculty of Medicine, Chulalongkorn University, Bangkok, Thailand; joined corresponding authors. Joined first authorships (Michael Maes, Asara Vasupanrajit).

## Abstract

**Background:** Major depression comprises two discrete subtypes, major (MDMD) and simple (SDMD) dysmood disorder. MDMD, but not SDMD, patients were identified to have highly sensitized cytokine/growth factor networks using stimulated whole blood cultures. However, no information regarding serum cytokines/chemokines/growth factors in SDMD is available.

**Objectives:** This case-control study compares 48 serum cytokines/chemokines/growth factors in academic students with SDMD (n=64) and first episode (FE)-SDMD (n=47) to those of control students (n=44) using a multiplex assay.

**Findings:** Both FE-SDMD and SDMD exhibit a notable inhibition of immune profiles, such as the compensatory immunoregulatory response system (CIRS) and alternative M2 macrophage and T helper-2 (Th-2) profiles. We observed a substantial reduction in the serum concentrations of five proteins: interleukin (IL)-4, IL-10, soluble IL-2 receptor (sIL-2R), IL-12p40, and macrophage colony-stimulating factor. A significant proportion of the variability observed in suicidal behaviors (26.7%) can be accounted for by serum IL-4, IL-10, and sIL-2R (all decreased), and CCL11 (eotaxin) and granulocyte CSF (both increased). The same biomarkers (except for IL-10), accounted for 25.5% of the variance in SDMS severity. A significant correlation exists between decreased levels of IL-4 and elevated ratings of the brooding type of rumination.

**Conclusions:** The immune profile of SDMD and FE-SDMD exhibits a significant deviation from that observed in MDMD, providing additional evidence that SDMD and MDMD represent distinct phenotypes. SDMD is characterized by the suppression of the CIRS profile, which signifies a disruption of immune homeostasis and tolerance, rather than the presence of an inflammatory response.

## Introduction

Research has provided evidence indicating that mood disorders are accompanied with the activation of the immune-inflammatory response (IRS) system (Maes et al., 1995a, Köhler et al., 2017). The activation of the IRS in mood disorders is accompanied by a mild inflammatory reaction, as evidenced by elevated levels of key proinflammatory cytokines, such as interleukin (IL)-1β and IL-6, and positive acute phase proteins, such hapto1globin, as well as complement factors (Maes, 1993). The immune-inflammatory theory of mood disorders was formulated by Maes et al. after doing research on individuals hospitalized with acute unipolar and bipolar depression (Maes et al., 1995a, Maes and Carvalho, 2018).

Maes et al. further developed this theory by including evidence suggesting both the IRS and the compensatory immune-regulatory system (CIRS) have a role in mood disorders (Maes et al., 1995b, Maes et al., 2012, Maes and Carvalho, 2018). The CIRS refers to the combined negative immune-regulatory and anti-inflammatory mechanisms that reduce the activity of the IRS and prevent excessive immune activation (Maes and Carvalho, 2018). The CIRS consists of immune-regulatory cytokines from the T helper-2 (Th-2) and T regulatory (Treg) lineages, specifically IL-4 and IL-10, as well as certain cytokine receptor levels, including soluble IL-2 receptor (sIL-2R) and soluble IL-1 receptor antagonist (sIL-1RA). Additionally, it includes acute phase proteins like haptoglobin (Maes et al., 1995b, Maes et al., 1997, Maes et al., 2012, Maes and Carvalho, 2018). During the acute phase of depression, many types of IRS cell lineages, such as M1 classical macrophage, Th-1, and Th-17 cells, become activated. However, there is a relative suppression of CIRS functions, resulting in a new setpoint where the IRS is more dominant than the CIRS (Maes and Carvalho, 2018). The IRS/CIRS theory of the acute stage of Major Depressive Disorder (MDD) suggests that heightened IRS activity and relatively weakened CIRS defenses are associated with elevated immune-associated neurotoxicity related to the immune system. This elevated neurotoxicity may lead to functional harm to neuronal and astroglial projections (Maes et al., 2012, Maes and Carvalho, 2018, Al-Hakeim et al., 2023).

A recent study utilizing unsupervised and supervised learning methods has identified two distinct clusters of patients within the sample of depressed individuals. These clusters are referred to as major dysmood disorder (MDMD) and simple dysmood disorder (SDMD) (Maes et al., 2023a). There are notable clinical distinctions between the two clusters, including heightened levels of depression, anxiety, fatigue, and lifetime and current suicidal behaviors (SBs) in MDMD. Additionally, patients with MDMD exhibit a higher index of reocurrence of illness (ROI) based on the number and frequency of suicidal behaviors, compared to those with SDMD. (Almulla et al., 2023, Michael et al., 2023). In addition, MDMD is distinguished by abnormalities in various pathways that are typically absent in patients with SDMD. These abnormalities include oxidative and nitrosative stress (O&NS), reduced antioxidant defenses, decreased levels of neurotrophic factors such as nerve growth factor, heightened atherogenicity, increased gut dysbiosis with a specific enterotype, heightened activation markers on T cells, and elevated levels of whole blood stimulated (LPS+PHA) in vitro production of cytokines, chemokines, and growth factors (Maes et al., 2022, Almulla et al., 2023, Maes et al., 2023a, Maes et al., 2023b). The latter findings indicate that MDMD is accompanied by activation of the JAK-STAT pathway as indicated by protein-protein interaction (PPI) analysis (Maes et al., 2022). The above pathway changes in MDMD are partly linked to the rise in ROI, suggesting that as the number of episodes and suicidal actions increases, the abnormalities in these pathways become more noticeable.

Maes et al. (2022) found no evidence of alterations in the whole blood-stimulated production of cytokines, chemokines, and growth factors (when challenged with LPS+PHA) in patients with SDMD. This suggests that unlike MDMD, there may be no heightened sensitivity in the immune network in this particular subtype of MDD. Therefore, it is crucial to analyze the immune condition in MDMD and SDMD individually, as well as investigate the immune pathways during the initial occurrence of these disorders (Maes et al., 2021).

Almulla et al. (2023) reported that FE-MDMD is characterized by significantly elevated serum IL-16 levels, heightened levels of several other serum cytokines and chemokines, indicating activation of M1 and Th-1 profiles, polarization towards Th-1 with activation of IRS, increased IRS/CIRS ratio. All in all, FE-MDMD seems to be primarily associated with the activation of IRS and, particularly, by Th-1 activation, which is additionally stimulated by elevated levels of many growth factors. However, there is a lack of data on the presence of cytokine/chemokine/growth factors in the serum of patients with SDMD and FE-SDMD (Almulla et al., 2023).

Therefore, this study aimed to analyze 48 serum cytokines/chemokines/growth factors using a multiplex assay in individuals with SDMD and FE-SDMD, in comparison to a control group. According to our prior research, we found no evidence of increased production of cytokines/chemokines/growth factors in SDMD when using LPS+PHA stimulated whole blood (Maes et al., 2022). Therefore, we would anticipate that there would be no indications of IRS activation in the serum of SMDM patients, contrasting with the activated IRS profiles discovered in MDMD.

## Subjects and methods

### Participants

In this case-control study, both depressed and nondepressed students participated. From November 2021 to February 2023, a total of 118 Thai-speaking university students of both gender and age groups (18–35) were recruited. The students were enrolled in various academic departments at Chulalongkorn University in Bangkok, Thailand. Interviews were administered to the depressed and normal control students at the Department of Psychiatry, King Chulalongkorn Memorial Hospital in Bangkok, Thailand. The depressed students met the criteria for MDD as outlined in the DSM-5 (American Psychiatric Association and Association, 2013). They also exhibited a Hamilton Depression Rating Scale (HAM-D) (Hamilton, 1960) score of at least 7, which excluded patients in remission, and a score below 22. Despite not meeting the diagnostic criteria for MDMD, every single patient was categorized as having SDMD. Forty-seven of the 64 patients with SDMD who were included were experiencing their initial depressive episode; therefore, they are referred to as first episode SDMD (FE-SDMD). Patients met the exclusion criteria if they had any of the following axis 1 and axis 2 psychiatric disorders: antisocial personality disorder, borderline personality disorder, substance use disorders, psycho-organic disorders, bipolar disorder, schizophrenia, schizoaffective disorder, anxiety disorders, autism spectrum disorders, or substance use disorders.

Forty-four healthy students were incorporated into the study if they had no documented lifetime history of axis-1 disorders (including dysthymia) or axis-2 disorders (antisocial and borderline personality disorders) or suicidal attempts, and their HAM-D score was below 7. Recruited via online advertisements and word-of-mouth, they were matched with patients according to their age, gender, and number of years of education. Participants who were at a heightened risk of suicidal ideation or who had neurological disorders including multiple sclerosis or any neuroinflammatory disorder were excluded from the study. Female students who were pregnant or lactating were not permitted to participate. Participants were not permitted to have any medical conditions that affected their immune system. These conditions included lupus erythematosus, inflammatory bowel disease, autoimmune disorders, rheumatoid arthritis, psoriasis, and inflammatory bowel disease. We also excluded students with cardiac, liver, endocrine and kidney disease, including hypo and hyperthyroidism, and chronic kidney disease. Students who had been diagnosed with moderate to severe COVID-19 infection or who had experienced modest COVID-19 symptoms in the month prior to their inclusion in the study were ultimately excluded. In addition, participants who exhibited potential long-lasting symptoms of COVID-19 (or any other infectious disease) such as persistent fatigue, congestion, and fever were ineligible. Three students were excluded from the study based on inclusion and exclusion criteria: two from the SDMD group and one from the control group.

The protocol for the study received approval from the Institutional Review Board (IRB) No.351/63 of the Faculty of Medicine, Chulalongkorn University in Bangkok, Thailand. Prior to the study, written informed consent was obtained from all participants.

### Clinical measurements

To gather socio-demographic information, a research assistant (AV) conducted a semi-structured interview. The interviewer collected the following: sex, age, years of education, relationship status, current smoking and alcohol use, family history of mental health conditions including major depressive disorder, bipolar disorder, anxiety disorders, psychosis, and suicide, and history of COVID-19 infection. The symptoms and severity of depression were assessed in this study utilizing the Beck Depression Inventory-II (BDI-II) (Beck et al., 1996) and the HAM-D (Hamilton, 1960). The self-rating questionnaire known as the BDI-II consists of twenty-one items that are evaluated on a four-point scale that spans from severe (3) to not present (0). (Mungpanich, 2008) translated the BDI-II into Thai, and this version possesses adequate validity and reliability. The Thai translation of HAM-D was utilized; it was authored by (Lotrakul et al., 1996).

The semi-structured interview known as the Columbia-Suicide Severity Rating Scale (C-SSRS) (Posner et al., 2011) was employed to assess both past (up to one month prior to participant inclusion) and present (within one month) suicidal behaviors (SB). The severity of suicidal ideation, the intensity of suicidal ideation, suicidal behaviors (including self-harm and attempts), and the lethality subscale comprise the four constructs of the C-SSRS. The source of the Thai translation was The Columbia Lighthouse Project (2016). By utilizing lifetime (LT), current suicidal ideation (SI), and suicidal attempt (SA) data, we have calculated a comprehensive score denoted as SB, which is an amalgamation of current + LT SB (Asara et al., 2023). Principal component analysis was employed to calculate the phenome score, as previously described, by utilizing the HAM-D, BDI-II, and SB scores as indicators. Rumination has been evaluated utilizing the Ruminative Response Scale (RRS) (Nolen-Hoeksema, 1991). The RRS consists of twenty-two items that are evaluated using a four-point scale that spans from almost never (1) to almost always (4). The Thai version (Thanoi and Klainin-Yobas, 2015) was utilized in this research, and it demonstrates satisfactory content validity and internal consistency (α = 0.90). The present study exclusively employs the brooding score from the rumination rating scale, as it exhibits a stronger correlation with depression scores than reflection. The first principal component derived from the brooding items of the RRS was designated as “brooding” (Asara et al., 2023). Given that brooding may account for a portion of the variance in the phenome of SDMD, we computed the residualized phenome scores through regression on the brooding scores. These scores represent the portion of the phenome that is not determined by brooding.

### Biomarker measurements

A total of 30 milliliters of fasting venous blood were collected from each student between 8:00 and 11:00 a.m. utilizing serum tubing and a disposable syringe. Following blood centrifugation at 35,000 rpm, serum was extracted and stored in Eppendorf containers containing small aliquots at -80 °C until they were defrosted in preparation for biomarker assays. We determined the concentrations and fluorescence intensities (FI) of forty-eight cytokines, chemokines, and growth factors using Bio-Plex Multiplex Immunoassay kits (Bio-Rad Laboratories Inc., Hercules, USA). Table 1 of the Electronic Supplementary File (ESF) contains a comprehensive inventory of the analytes utilized in our research, including their corresponding gene IDs and aliases. In summary, the methodology comprised the subsequent stages: a) The serums were diluted with sample diluent (H.B.) (1:4). b) The diluted samples were added to a 96-well plate containing 50µl of microparticle cocktail (including cytokines/chemokines/growth factors) per well. The plate was then incubated at room temperature with 850 rpm shaking for one hour. c) Following three rinses, 50µl of diluted Streptavidin-PE was added to each well. The plate was then agitated at room temperature for ten minutes. d) Following this, the sample was transferred to the 96-well plate. The Bio-Plex® 200 System (Bio-Rad Laboratories, Inc.) was utilized in the subsequent analysis to evaluate the cytokines/chemokines/growth factors. The intra-assay CV values for every analyte were found to be below 11.0%. The concentrations were determined using the manufacturer-supplied standard concentrations. The percentage of concentrations exceeding the lowest measurable concentration (OOR) was subsequently assessed. Table 1 provides a list of the measurable analytes in ESF, along with the proportion of values that exceeded the OORs. FI values are more appropriate than absolute concentrations (especially when numerous plates are utilized), and, therefore, our statistical analysis utilizes blank subtracted FI values. Values less than OOR values were approximated by employing the sensitivity of the assays. Our statistical analysis performed on single cytokines/chemokines/growth factors excluded analytes with concentrations that were out of range in more than 50% of the instances. In the regression analysis, variables that possessed quantifiable levels ranging from 10% to less than 50% were regarded as prevalences, or dummy variables. As a result, VEGF and FGF were transformed into dummy variables for the purpose of conducting the analysis. The variables used to construct the following ESF profiles are presented in Table 2: M1, M2, z M1 – z M2 (reflecting M1 polarization), Th-1, Th-2, z Th-1 – z Th-2 (reflecting Th-1 polarization), IRS, CIRS, and z IRS – z CIRS (reflecting the IRS/CIRS ratio).

**Table 1.** Socio-demographic and clinical data of students with simple dysmood disorder (SDMD) and control students (HC).

| **Biomarkers** | **HC (n=44)** | **SDMD (n=64)** | **F/X<sup>2</sup>** | **df** | **p** |
| --- | --- | --- | --- | --- | --- |
| Age (years) | 23.5 (3.2) | 22.4 (3.2) | 2.75 | 1/106 | 0.100 |
| Sex (F/M) | 37/7 | 54/10 | 0.00 | 1 | 0.968 |
| BMI (kg/m<sup>2</sup>) | 22.3 (3.6) | 22.4 (5.1) | 0.01 | 1/106 | 0.927 |
| Education (years) | 16.6 (1.5) | 15.9 (2.7) | 2.15 | 1/106 | 0.146 |
| Duration (range) | - | 1 – 108 months | - | - | - |
| Single/relation or marriage | 27/17 | 27/37 | 3.84 | 1 | 0.050 |
| Smoking (No/Yes) | 43/1 | 59/5 | 1.53 | 1 | 0.217 |
| Student drinking (No/Yes) | 41/3 | 53/11 | 2.49 | 1 | 0.115 |
| Suffered from mild COVID-19 (No/Yes) | 30/14 | 49/15 | 0.93 | 1 | 0.334 |
| Total HAM-D score | 2.4 (2.0) | 14.6 (4.7) | 257.60 | 1/106 | <0.001 |
| Total BDI-II score | 6.7 (5.4) | 24.7 (11.5) | 86.85 | 1/106 | <0.001 |
| SB score (z score) | -0.872 (0.199) | 0.599 (0.881) | 118.13 | 1/106 | <0.001 |
| Phenome (z score) | -0.998 (0.300) | 0.686 (0.682) | 236.91 | 1/106 | <0.001 |
| Brooding (z score) | -0.802 (0.809) | 0.551 (0.702) | 85.57 | 1/106 | <0.001 |
| Residualized phenome (z score) | -0.716 (0.091) | 0.482 (0.071) | 85.82 | 1/105 | <0.005 |

All data are shown as mean (SD) or as ratios. F: results of analysis of variance; X<sup>2</sup>: results of contingency analysis.
HAM-D: Hamilton Depression Rating Scale; BDI-II: Beck Depression Inventory; SB: suicidal behaviors; Residualized phenome: brooding subtracted phenome scores (following regression analysis)

**Table 2.** Differences in immune profiles between students with simple dysmood disorder (SDMD) and healthy students (HC).

| **Biomarkers (z scores)** | **HC** | **SDMD** | **F** | **df** | **P** |
| --- | --- | --- | --- | --- | --- |
| All immune profiles* |  |  |  |  |  |
| IRS | 0.159 (0.150) | -0.111 (0.125) | 1.88 | 1/103 | 0.174 |
| **CIRS** | **0.455 (0.142)** | **-0.304 (0.118)** | **16.67** | **1/103** | **<0.001** |
| **zIRS-zCIRS** | **-0.245 (0.126)** | **0.171 (0.105)** | **6.34** | **1/103** | **0.013** |
| M1 | 0.131 (-0.092) | -0.092 (0.127) | 1.24 | 1/103 | 0.268 |
| **M2** | **0.231 (0.149)** | **-0.161 (0.124)** | **4.04** | **1/103** | **0.047** |
| zM1-zM2 | -0.100 (0.154) | 0.070 (0.128) | 0.71 | 1/103 | 0.402 |
| **Th1** | **0.266 (0.147)** | **-0.186 (0.122)** | **5.50** | **1/103** | **0.021** |
| **Th2** | **0.314 (0.143)** | **-0.219 (0.119)** | **8.10** | **1/103** | **0.005** |
| zTh1-zTh2 | -0.048 (0.143) | 0.033 (0.120) | 0.18 | 1/103 | 0.666 |
| The significant cytokines* |  |  |  |  |  |
| **sIL-2R** | **0.243 (0.147)** | **-0.167 (0.122)** | **5.92** | **1/101** | **0.017** |
| **IL-4** | **0.337 (0.143)** | **-0.231 (0.118)** | **9.14** | **1/101** | **0.003** |
| **IL-10** | **0.297 (0.146)** | **-0.204 (0.121)** | **8.10** | **1/101** | **0.005** |
| **IL-12p40** | **0.347 (0.143)** | **-0.238 (0.118)** | **9.74** | **1/101** | **0.002** |
| **M-CSF** | **0.320 (0.146)** | **-0.220 (0.121)** | **7.52** | **1/101** | **0.007** |

\*Data were adjusted for age, body mass index, sex, smoking and alcohol use.
IRS: immune-inflammatory response system; CIRS: compensatory immune-regulatory system; M: macrophage; Th: T helper; IL\_interleukin, sIL-2R: soluble IL-2 receptor; M-CSF: macrophage colony stimulating factor.

### Statistical analyses

Statistical associations between categorical variables were assessed utilizing a contingency table analysis (X^2^-test). The researchers employed analysis of variance (ANOVA) to examine the relationships between diagnostic categories and clinical data. The analysis examined the correlations between continuous variables by employing Pearson’s product-moment correlation coefficients or Spearman’s rank order correlation coefficients. Manual and step-up automated multiple regression analysis were implemented to delineate the most important predictors (biomarkers) of the dependent variables (the clinical data). The automated method was performed with p-values of 0.05 and 0.06 as the entry and exclusion criteria, respectively. The essential metrics of the model were calculated, namely the total variance explained (R^2^, used as effect size), F, df, and p-values, as well as the standardized beta coefficients with t and p values for the separate predictors. Homoskedasticity was established by employing the White and modified Breusch-Pagan tests. The variance inflation factor (VIF) and tolerance were used to check for collinearity issues. In addition, the condition index and variance proportions were assessed to obtain a more comprehensive understanding of multicollinearity. Residuals, residual plots, and data quality were consistently assessed in the final model. We conducted partial regression analyses, which involved generating partial regression graphs, in addition to the linear modeling analyses. An automatic stepwise binary logistic regression analysis was performed to delineate the best prediction of SDMD or FE-SDMD versus controls. Univariate generalized linear models (GLMs) were employed to assess the associations between the diagnosis of SDMD and FE-SDMD and immune profiles as well as isolated cytokines/chemokines/growth factors. Statistical significance was determined using a p-value of 0.05 and two-tailed tests.

The primary statistical analysis employed in this study was multiple regression analysis, which examined the relationship between the phenome data (dependent variable) and the immunological markers (predictors). Due to the number of participants (n=108), the number of predictors was constrained to five. Based on an effect size of 0.176, which accounts for approximately 15% of the variance, together with the inclusion of 5 covariates, a desired statistical power of 0.08, and a significance level of 0.05, the minimum required sample size is determined to be 79. This calculation was performed using G*Power 3.1.9.7 software.

## Results

### Sociodemographic and medical information

**Table 1** contains information regarding sociodemographics. SDMD students and control students did not differ significantly about age, gender, BMI, years of education, marital status, smoking or alcohol use, or a prior diagnosis of mild COVID-19. SDMD patients exhibited substantially higher scores on the HAM-D, BDI-II, SB, phenome, brooding, and residualized phenome (after regression on brooding) scores as compared to the healthy control group.

### Immunological assessments in SDMD and FE-SDMD

The immune profile differences between patients with SDMD and healthy controls (HC) are presented in **Table 2**. In SDMD, the CIRS profile was considerably diminished in comparison to the control group, whereas no substantial disparities were observed in the IRS profile. This explains why the IRS/CIRS ratio in patients was significantly greater than in controls. Patients had diminished M2, Th-1, and especially Th-2 profiles compared to the control group. A univariate GLM analysis was conducted on the individual biomarkers and identified variations in five of them: sIL-2R, IL-4, IL-10, IL-12p40, and M-CSF. Some patients were treated with sertraline (n=20), fluoxetine (n=10), venlafaxine (n=7), escitalopram (n=16), benzodiazepines (n=29), and atypical antipsychotics (n=12) and, therefore, we have statistically adjusted for the drug state of the patients. We could not find any significant effects of sertraline (p=0.056), fluoxetine (p=0.153), venlafaxine (p=0.402), escitalopram (p=0.878), benzodiazepines (p=0.245), and atypical antipsychotics (p=0.121) on the immune profiles, while all effects of SDMD remained significant. Likewise, we could not find any significant effects of sertraline (p=0.159), fluoxetine (p=0.556), venlafaxine (p=0.111), escitalopram (p=0.870), benzodiazepines (p=0.771), and atypical antipsychotics (p=0.081) on sIL-2R, IL-4, IL-10, IL-12p40, and M-CSF.

ESF, Table 3 presents the immune profiles and cytokines/growth factors in FE-SDMD as opposed to controls. FE-SDMD was distinguished by a decreased CIRS, an elevated IRS/CIRS ratio, and a decreased Th-2 profile. ESF, Table 4 shows that IL-4, IL-10, and II-12p40 were significantly decreased in FE-SDMD relative to controls, whereas there was a trend towards lower sIL-2R and M-CSF in FE-SDMD relative to controls (p=0.055), a finding that was statistically significant using a one-tailed test.

**Table 3.** Results of a binary logistic regression analysis with first-episode simple dysmood disorder as dependent variable (controls as reference group).

| **Explanatory variables** | **B** | **SE** | **Wald (df=1)** | **P** | **Odds ratio** | **Lower 95% limit** | **Upper 95% limit** |
| --- | --- | --- | --- | --- | --- | --- | --- |
| CIRS | -0.918 | 0.474 | 7.59 | 0.006 | 0.399 | 0.208 | 0.767 |
| IL-4 | -1.448 | 0.296 | 9.32 | 0.002 | 0.235 | 0.093 | 0.596 |
| G-CSF | 1.042 | 0.416 | 12.42 | <0.001 | 2.835 | 1.588 | 5.062 |
| CCL11 | 1.019 | 0.245 | 5.99 | 0.014 | 2.771 | 1.225 | 6.268 |

CIRS: compensatory immune-regulatory system; CCL11 or eotaxin, IL: interleukin, G-CSF: granulocyte colony stimulating factor.

**Table 4.** Intercorrrelation matrix (Pearson’s correlation coefficients) between clinical scores and immune profiles in simple dysmood disorder.

| Variables (z scores) | HAM-D | BDI-II | Brooding | SB | Phenome |
| --- | --- | --- | --- | --- | --- |
| IRS | -0.202* | -0.182 | -0.141 | -0.177 | -0.208* |
| CIRS | -0.310*** | -0.287** | -0.275** | -0.292** | -0.330*** |
| zIRS-zCIRS | 0.128 | 0.124 | 0.159 | 0.135 | 0.143 |
| M1 | -0.129 | -0.108 | -0.106 | -0.160 | -0.147 |
| M2 | -0.240* | -0.189 | 0.190* | -0.204* | -0.234** |
| zM1-zM2 | 0.111 | 0.080 | 0.084 | 0.044 | 0.087 |
| Th1 | -0.232* | -0.300** | -0.256** | -0.315*** | -0.314*** |
| Th2 | -0.367*** | -0.284** | -0.327*** | -0.322*** | -0.361*** |
| zTh1-zTh2 | 0.140 | -0.017 | 0.073 | 0.007 | 0.048 |

\* $p < 0.05$ ; \*\* $p < 0.01$ ; \*\*\* $p < 0.001$
HAM-D: Hamilton Depression rating scale score; BDI-II: Beck Depression Inventory score; SB: suicidal behavior scores; IRS: immune-inflammatory response system; CIRS: compensatory immune-regulatory system; M: macrophage; zM1-zM2: index for classical versus alternative M polarization; Th: T helper; zTh1-zTh2: index for Th1 versus Th2 polarization; IL: interleukin.

The outcome of a binary logistic regression analysis utilizing the biomarkers to differentiate FE-SDMD from controls is presented in **Table 3**. A significant differentiation between the two groups was achieved by employing 4 immune markers (X^2^=35.52, df=4, p<0.001, Nagelkerke=0.381): CIRS and IL-4 (both inversely associated), and CCL11 and G-CSF (both positively associated). As indicated by the confusion matrix, 71.0% of the subjects were classified accurately, with a specificity of 61.4% and a sensitivity of 77.8% (Nagelkerke=0.407).

### Intercorrelations between immune and clinical data

The intercorrelation matrix between clinical (HAM-D, BDI-II, brooding, SB, and phenome) and immune profiles is presented in **Table 4**. IRS was inversely and significantly correlated with HAM-D and phenome scores. All five clinical data exhibited an inverse association with the M2 immune profile, except for the BDI-II-score. There was a significant negative correlation observed between CIRS, Th-1, and Th-2 and all five clinical rating scale scores. The partial regression of the phenome score on CIRS values (adjusted for age, sex and education) is depicted in **Figure 1**.

**Figure 1.**
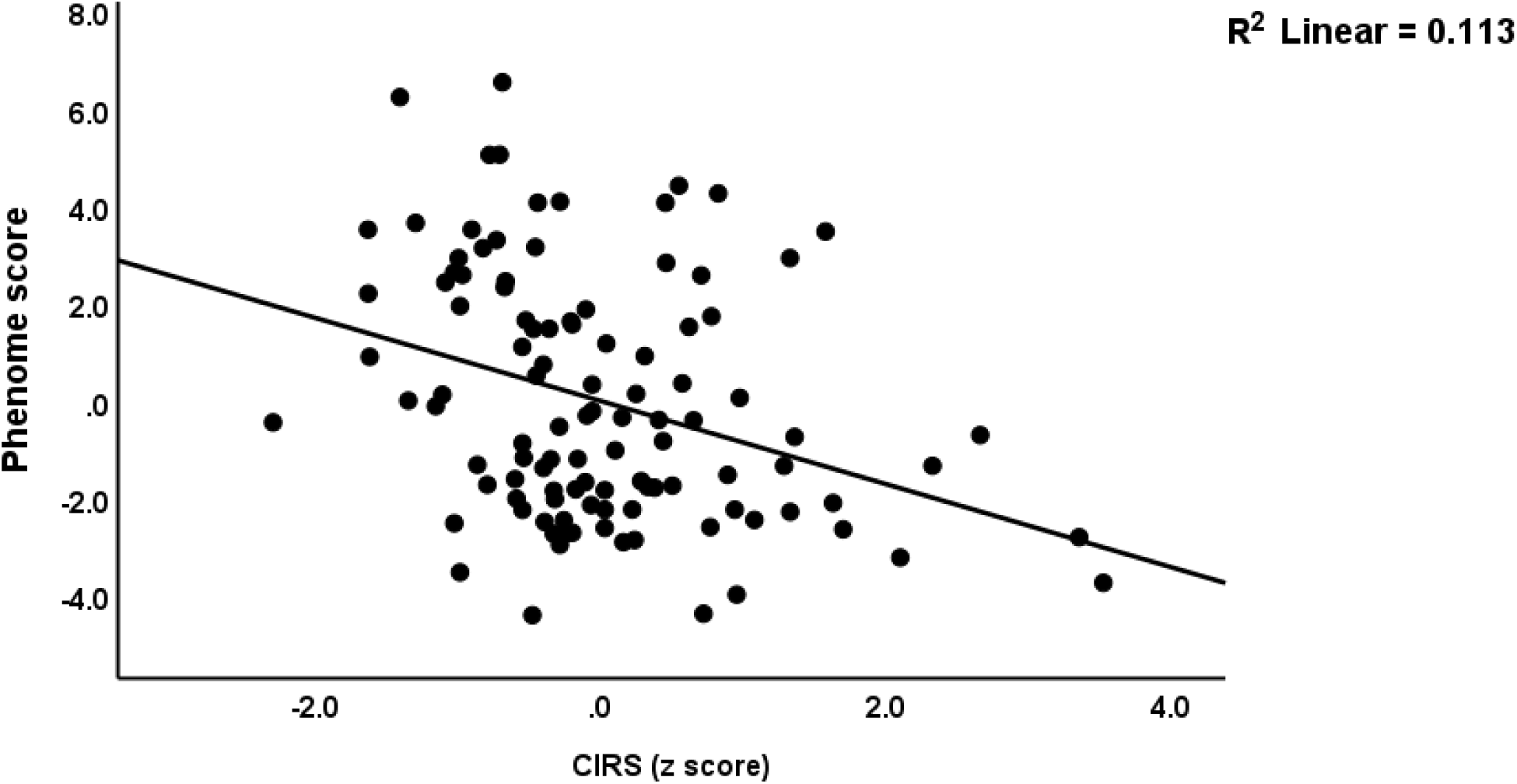
Partial regression of the phenome score on the compensatory immunoregulatory system (CIRS) score in patients with simple dysmood disorder (p<0.001).

### Findings from multiple regression analysis

The outcomes of regression analyses utilizing biomarkers as explanatory variables and clinical data as dependent variables are presented in **Table 5**. We utilized two distinct sets of explanatory variables: a) immune biomarkers alone coupled with demographic data; and b) the immune biomarkers, demographic data, and brooding. Therefore, the latter regression calculates the impact of the biomarkers on that part of the phenome that is unaffected by brooding. 26.7% of the variance in SB is accounted for by IL-4, sIL-2R, and IL-10 (all three inversely), and by G-SCF and CCL11 (both positively), as shown in Regression 1a. **Figure 2** shows this partial regression plot of SB on IL-4 levels. By adjusting for the influences of brooding in regression 1b, it is observed that 49.2% of the variability in SB can be accounted for by brooding and G-SCF (both positively), and by IL-4, PDGF, and IL-10 (all three inversely). 24.4% of the variance in HAM-D is accounted for (negatively) by IL-4 and (positively) by CCL11 and G-CSF, according to regression 2a. Regression 2b reveals that a significantly greater proportion of the variance (62.6%) is accounted for by brooding, SDF, and CCL11 (all three in a positive manner), whereas IL-4 and M-CSF are inversely correlated. A portion of the variance in the BDI-II score (17.2%) could be accounted for by IL-18 (increased) and IL-4 and SCGF (inversely), whereas 61.9% could be accounted for by the brooding and IL-1α (both positively) and SCGF (negatively). Regression analysis #4a revealed that 25.5% of the variance in the phenome score could be accounted for by G-CSF and CCL11 (positively) and IL-4 and sIL-2R (negatively). The partial regression of the phenome score on IL-4 is illustrated in **Figure 3**. According to regression 4b, 65.4% of the variance in the phenome score was accounted for by brooding, and CCL11 (both in a positive manner), while IL-4 and age were found to be inversely related. Regression #5 shows that 13.9% of the variance in brooding was explained by education and IL-4 (both inversely).

**Figure 2.**
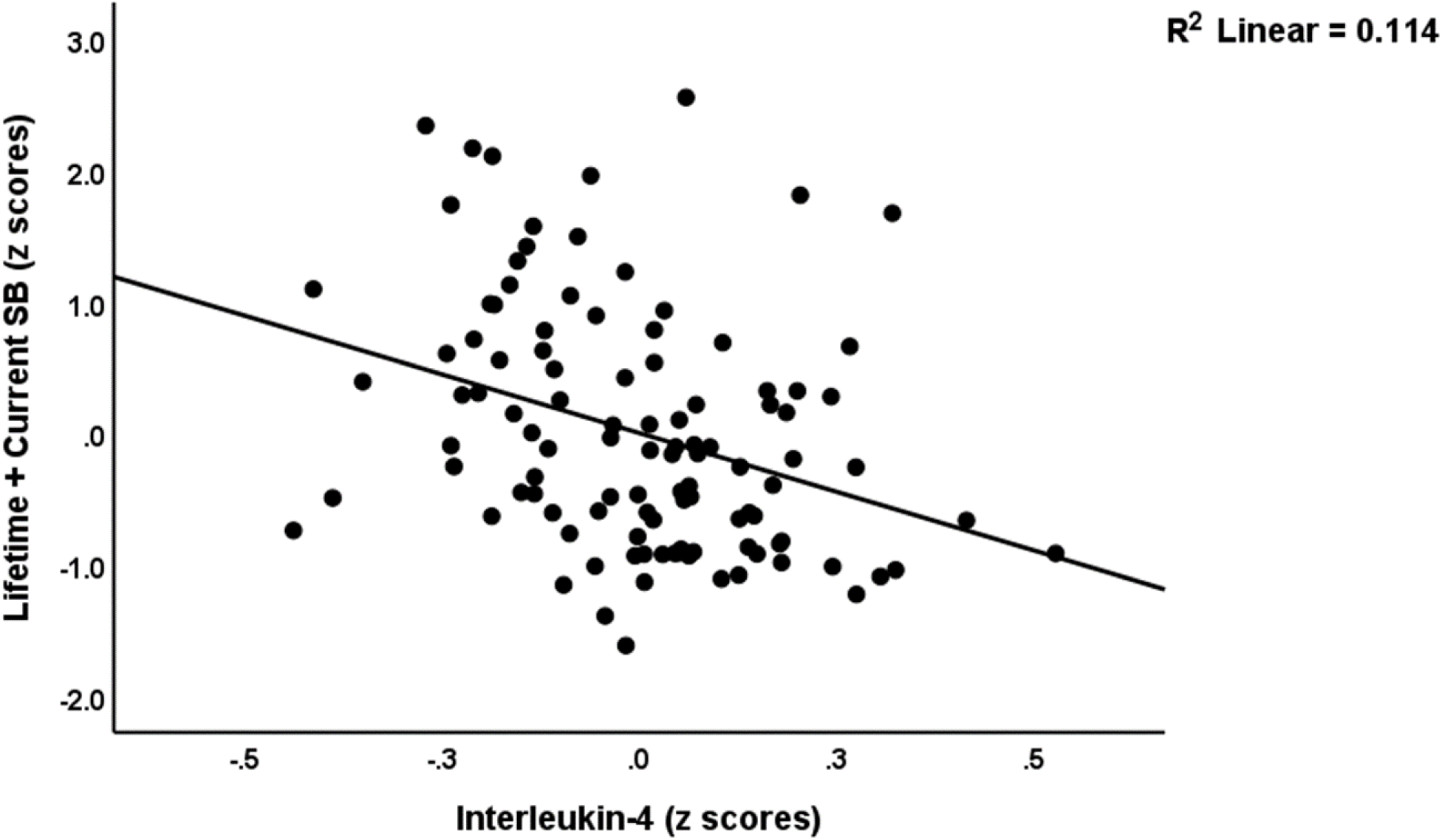
Partial regression plot of lifetime and current suicidal behaviors (SB) on interleukin-4 levels in patients with simple dysmood disorder (p<0.001).

**Figure 3.**
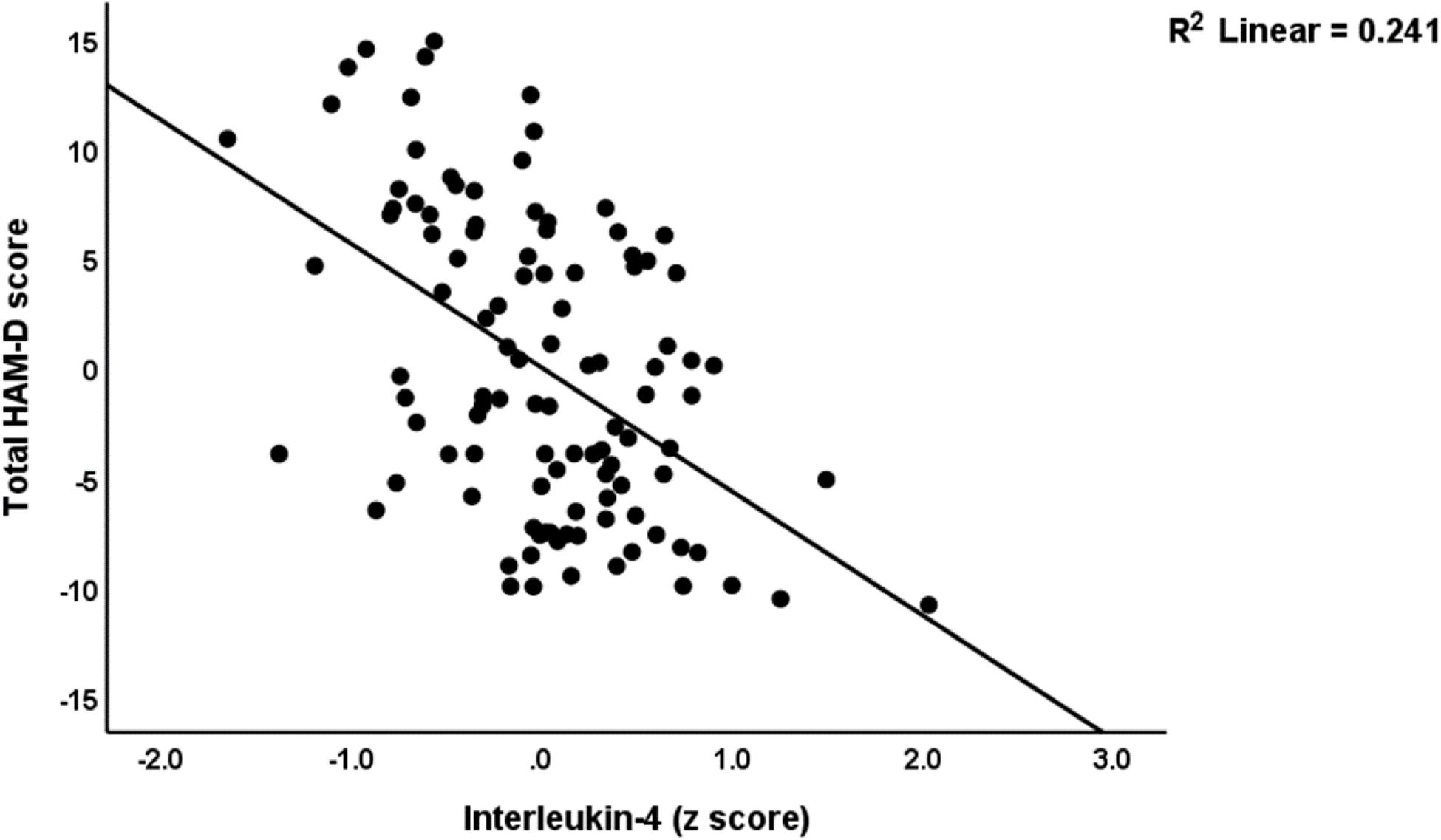
Partial regression of the Hamilton Depression rating Scale (HAM-D) score on serum interleukin-4 in patients with simple dysmood disorder (p<0.001).

**Table 5.**
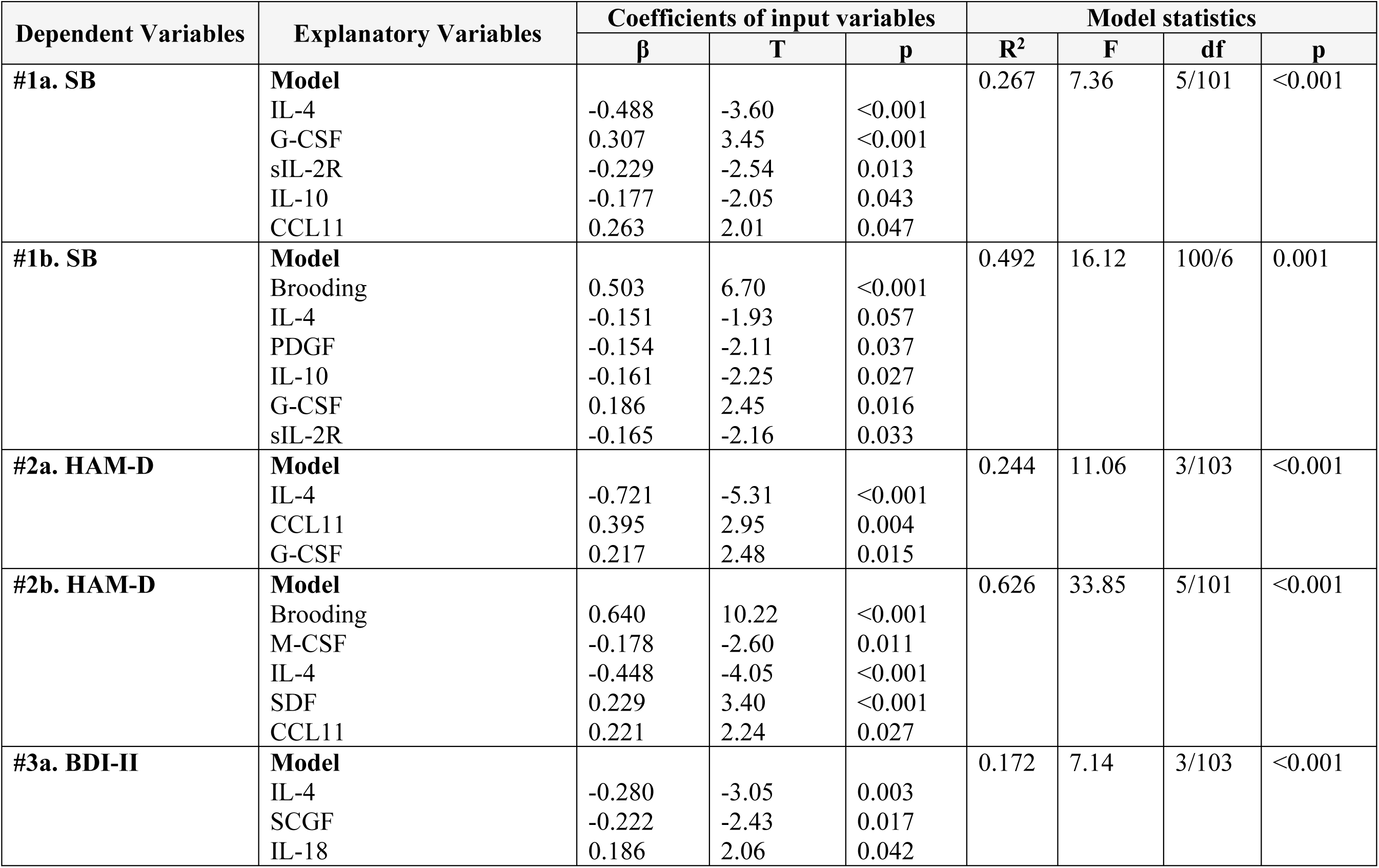
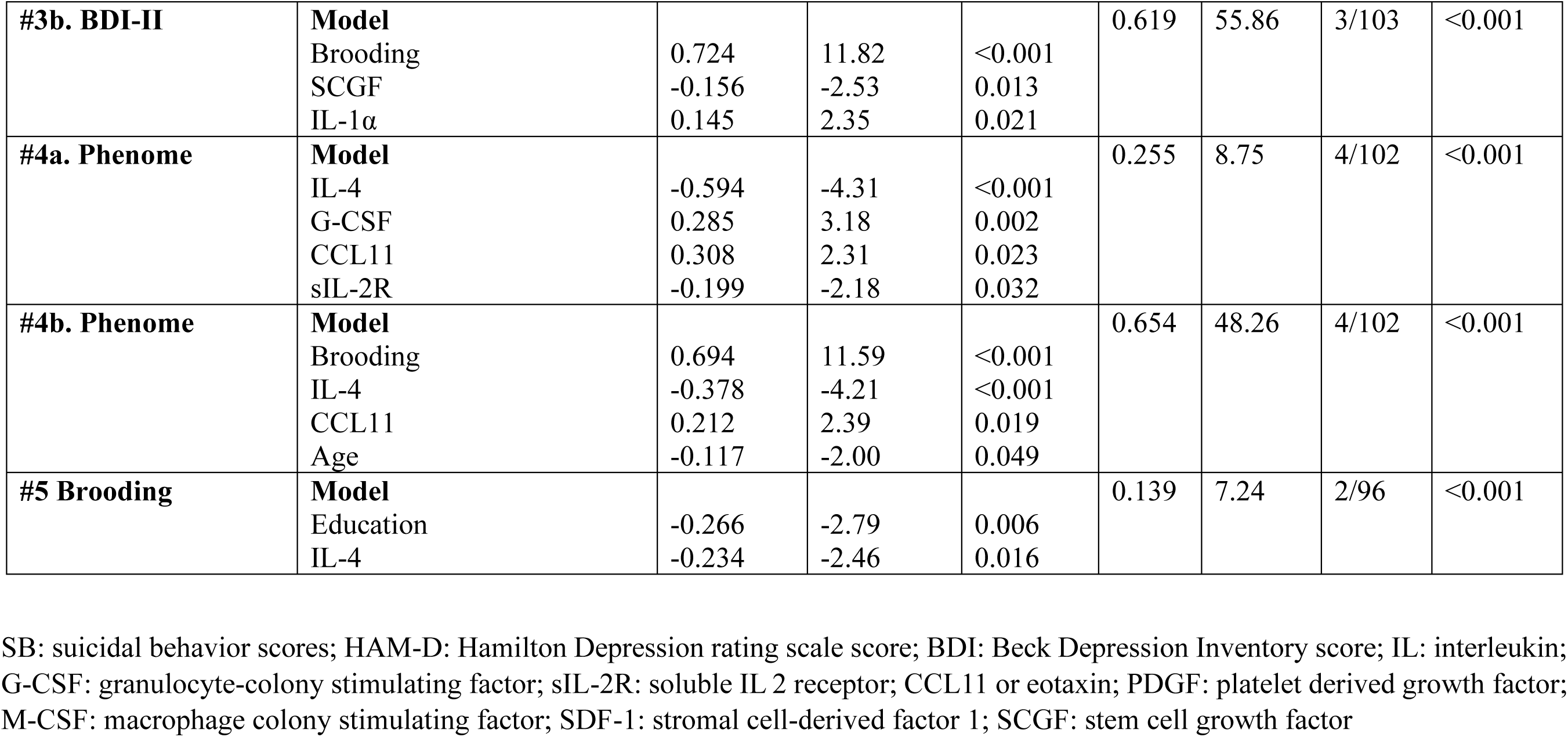
Results of multiple regression analyses with suicidal behaviors or clinical scores as dependent variables and immune biomarkers as explanatory variables.

## Discussion

### Attenuated CIRS in FE-SDMD

The first significant discovery of the current research is that SDMD and FE-SDMD are associated with substantial suppression of immune profiles, including M2 and Th2 profiles, and the comprehensive CIRS profile, in particular. This attenuated CIRS profile in the SDMD outpatients differs significantly from the profile observed in patients with FE-MDMD, acute major depression, and acutely hospitalized depressed patients. Acute episodes of major depression in hospitalized patients are invariably and conspicuously accompanied by immune activation, including Th-1, Th-2, and Th-17 activation, as discussed in the introduction (Maes and Carvalho, 2018). However, there are indications that the CIRS is activated to a lesser extent than the IRS during the acute phase of MDD in hospitalized patients, resulting in a moderately elevated IRS/CIRS ratio (Maes and Carvalho, 2018). Similarly, during the acute phase of severe depression, there is an elevation in the T effector to T regulatory cell ratio (Michael et al., 2023). In addition, the established profile in FE-MDMD differs significantly from that of FE-SDMD. The former reveals increased chemokine levels, a powerfully activated growth factor network, T cell activation, Th-1 activation, and Th-1 polarization, in addition to M1 activation (Almulla et al., 2023). However, the current findings align with those of Maes et al. (2022), who failed to identify any substantial immune network sensitization in SDMD but observed a robust sensitization in MDMD (owing to the stimulation of whole blood production by LPS and PHA).

It is important to highlight that for the purpose of analyzing the immune profile of SDMD and FE-SDMD, we opted to utilize a homogeneous sample of outpatients consisting of students exhibiting a mild manifestation of MDD. Therefore, in contrast to conventional approaches to depression research, which involve exclusively inpatients, mixed samples of mild and severely depressed patients, or mixed samples of MDD and SDMD patients combined with individuals in remission or partial remission, our study sample comprises SDMD outpatients only. Hence, the present results support our view that to investigate the pathogenesis of depression, it is necessary to partition the MDD subgroup into its two distinct subsamples: SDMD and MDMD (Maes et al., 2023b).

Certain researchers may consider the MDMD subgroup to be “inflammation-associated depression,” a subtype of depression distinguished by the presence of “inflammation,” while the SDMD subgroup is devoid of “inflammation.” All we can do is hope that our results are not erroneously interpreted that way. While patients with acute MDMD do exhibit a mild inflammatory response, which is characterized by elevated acute phase proteins and complement factors, and an activated M1 profile (Maes, 1993), the term “IRS activation” is more appropriate to denote the immune disorders in MDMD. IRS activation, in fact, is the umbrella term that encompasses both inflammation and the activation of other profiles (including M1, M2, Th-1, Th-2, Th-17 and T regulatory). Furthermore, it is important to emphasize that throughout the partial remission phase, the immune system may have undergone such a transformation that the CIRS activities have become more active than the IRS, which could potentially signify a state of repair and homeostasis (Maes et al., 2021). Significantly more notably, SDMD is characterized by a distinct immune profile — a suppressed CIRS profile and a relative increase in immune compounds with known neurotoxic effects — as will be elaborated upon in the following sections.

### Role of the affected CIRS components in SDMD

The results of our secondary analyses, which were conducted on the distinct cytokines/chemokines/growth factors, indicated a significant decrease in the serum levels of five proteins: IL-4, IL-10, sIL-2R, IL-12p40, and M-CSF. Thus, CIRS-associated proteins or inhibitors of pro-inflammatory pathways comprise four of the five proteins that exhibited a decrease in SDMD: IL-4, IL-10, sIL-2R, and IL-12p40. Once more, these results exhibit a significant divergence from the acute phase of severe mood disorders, during which elevated concentrations of sIL-2R, IL-4, and IL-10 may be observed (Maes et al., 1990, Maes and Carvalho, 2018, Al-Fadhel et al., 2019, Al-Hakeim et al., 2020). IL-4 levels may be elevated in patients with bipolar disorder, according to two recent meta-analyses (Modabbernia et al., 2013, Munkholm et al., 2013). In comparison to the control group, IL-10 is substantially elevated in patients with major depression, according to a recent meta-analysis (Köhler et al., 2017). However, despite the overall activation of immune profiles that accompanies MDMD, neither IL-4 nor IL-10 levels increase significantly, suggesting a relative deficiency in these crucial immunoregulatory cytokines (Almulla et al., 2023).

IL-4 is recognized for its negative immunoregulatory and anti-inflammatory properties. It hinders the development of Th-1 macrophages, promotes Th-2 polarization, and can also stimulate M2 macrophage activation (Maes and Carvalho, 2018). Furthermore, in addition to inhibiting the synthesis of TNF-α and IL-6, this cytokine stimulates the secretion of IL-10, TGF-β, and sIL-1RA, among other anti-inflammatory substances (Woodward et al., 2010). Additionally, IL-4 regulates the JAK-STAT pathway and regulates the aryl hydrocarbon receptor (AHR), which is involved in immune homeostasis and T regulatory functions (Shinde and McGaha, 2018, Rothhammer and Quintana, 2019, Deimel et al., 2021). Like IL-4, IL-10 functions as a significant immunosuppressive, anti-inflammatory, and immunoregulatory cytokine. It has the potential to stimulate the differentiation of T0 cells into induced T regulatory cells. Additionally, Th-1 cytokines, including IL-12 and IFN-γ, and Th-1, Th-2, and Th-17 cells are inhibited by IL-10, thereby preventing hyperinflammation by acting as an immunosuppressor (Romagnani et al., 2000, Moore et al., 2001, Mosser and Zhang, 2008, Vignali et al., 2008, Chapoval et al., 2010, Tran, 2012, Adeegbe and Nishikawa, 2013). Critically, immune homeostasis and tolerance is maintained and regulated by T regulatory cells and IL-10 (Vignali et al., 2008).

Despite serving as a surrogate indicator for T cell activation, elevated sIL-2R levels inhibit proliferation, natural killer cell functions, and cytotoxicity that are all induced by IL-2 (Dummer et al., 1992). A plausible mechanism posits that sIL-2Rs establish a complex with plasma IL-2, sIL-2R-IL-2, which inhibits IL-2 availability for IL-2 signaling and stimulates T regulatory cell differentiation (Maes et al., 1991, Caruso et al., 1993, Witkowska, 2005, Yang et al., 2011, Vanmaris and Rijkers, 2017). Zagozdzon and Lasek (2016) suggest that IL-12p40 could impede IL-12 signaling via IL-12p70, thus interfering with IL-12 and, by extension, Th-1-associated mechanisms. By reducing the concentrations of these four cytokines (or their receptors), the negative feedback on M1, Th-1, Th-17, IL-2, and IL-12p70 signaling may be mitigated. Consequently, decreased concentrations of these 4 proteins in individuals with SDMD may contribute to compromised immune tolerance and compromised immune homeostasis, ultimately resulting in impaired immunoregulation (Zagozdzon and Lasek, 2016).

It is noteworthy to mention that SDMD is also associated with decreased concentrations of serum M-CSF. Microglia and macrophages may become polarized toward the alternative M2-like phenotype in response to this growth factor (Pons and Rivest, 2018). Therefore, by inhibiting the release of proinflammatory mediators, M-CSF potentially facilitates the process of tissue repair. It can be deduced that the diminished M-CSF observed in SDMD may have suppressed this negative feedback signaling, thus playing a role in the heightened toxicity. The subsequent section delves into the consequences of the decreased CIRS profile and CIRS proteins.

### Increased immune-associated neurotoxicity in SDMD

The third significant discovery of this research is that the decreased CIRS profile and specific anti-inflammatory cytokines are linked to all essential phenotypic characteristics of SDMD, such as self-rating scales or interview-based depression severity, suicidal tendencies, and ruminating. During our regression analyses, IL-4 consistently emerged as the most reliable predictor of the key phenome characteristics, with sIL-2R ranking as the second most significant factor. However, after accounting for the influence of CIRS, IL-4, and sIL-2R, we noted that certain cytokines/chemokines/growth factors exhibited a positive correlation with the aforementioned critical characteristics of SDMD or with the diagnosis of SDMD in comparison to the control group.

CCL11 was the most significant positive predictor of the phenome characteristics, followed by G-CSF. It is crucial to emphasize that SDMD does not inherently result in elevated levels of CCL11 or GM-CSF. However, when the effects of decreased CIRS components are considered, a significant association is observed with both proteins. Therefore, it is possible that CCL11 and GM-CSF could maybe induce some toxic effects, solely due to a reduction in CIRS protection and a breakdown in immune tolerance.

In the past, research on CCL11 in major depression yielded inconsistent findings, as evidenced by both positive and negative results (Grassi-Oliveira et al., 2012, Magalhaes et al., 2014, Teixeira et al., 2018, Al-Hakeim et al., 2020, Almulla et al., 2023). Furthermore, CCL11 was not substantially elevated in depression, according to a recent meta-analysis (Leighton et al., 2018). CCL11 is a neurotoxic chemokine that has the potential to traverse the blood-brain barrier, as was previously mentioned (Michelle et al., 2014). Chemotaxis is regulated by this chemokine via its interaction with CCR3. It is also implicated in allergic airway diseases, inflammatory bowel disease, and gastro-intestinal allergic hypersensitivity (Amerio et al., 2003). By increasing oxidative stress, inhibiting neurogenesis, and interfering with the blood-brain barrier, CCL11 may have neurotoxic effects. (Jamaluddin et al., 2009, Parajuli et al., 2015). In the past, no substantial alterations in G-CSF levels were observed in patients diagnosed with MDMD (Almulla et al., 2023). While G-CSF has been associated with potential long-term neuroprotective effects (Song et al., 2016), it may exert detrimental effects on inflammation and neutrophil-mediated immunity (Song et al., 2016, Martin et al., 2021).

Naturally, the inquiry pertains to whether this proportional elevation in serum CCL11 and G-SCF could potentially influence brain functions in any way. Nevertheless, the consequences of decreased CIRS components are more evident, as they have a direct influence on the overall functions of CIRS and T regulation and may also have additional immune and generalized effects. As an illustration, reduced concentrations of IL-4 have the potential to impact the JAK-STAT pathways (Deimel et al., 2021). Conversely, decreased levels of IL-10 may disrupt the IL-10/STAT3 pathway, which is critical for immune tolerance and T regulatory functions (Schmetterer and Pickl, 2017). As previously mentioned, IL-4 controls the activity of the AHR, which has numerous physiological and biochemical effects, including antioxidant, detoxification, and immune functions. Additionally, it influences microbial defenses, energy metabolism, xenobiotic metabolism, organ homeostasis (including that of the nervous and cardiac systems), and IL-10 production (Tanaka et al., 2005, Neavin et al., 2018, Rothhammer and Quintana, 2019, Barroso et al., 2021, Granados et al., 2022). In addition to inducing the AHR via a STAT6-related pathway, IL-4 may also promote the AHR’s translocation to the nucleus (Tanaka et al., 2005).

### IL-4 and brooding

Given that ruminating over a variety of factors, including recent life events, accounts for a greater proportion of the variance in the SDMD phenome (Asara et al., 2023), we have adjusted our statistical analyses to exclude the brooding-explained variance. As described in the Results section, the inclusion of brooding as an additional explanatory variable had no impact on the outcomes. This suggests that the manifestation of depression, as assessed by self-rating (BDI-II) and psychiatric interview (HAM-D) scores, comprises at least two distinct components: one, which can be elucidated through pondering and rumination, and the other, which is contingent upon aberrations in the immune system. Thus, depressive phenomenology in patients with SDMD and FE-SDMD is likely explained by a combination of cognitive processes, including rumination, and immune pathways, according to these findings. In addition, IL-4 accounts for a proportion of the variance in brooding. This is particularly critical, given that ruminating is a symptomatic manifestation of depression and is a manifestation of the same factor as HAM-D, BDI-II, and SBs (Asara et al., 2023). Hence, it can be deduced that immune pathways play a critical role in determining the various manifestations of the phenome of depression assessed here, with the additional factor being brooding’s association with reduced IL-4.

### Limitations

If additional biomarkers of MDD had been assessed, such as neurotrophic compounds (e.g., brain-derived neurotrophic brain markers), astroglial and neuronal injury markers (e.g., neurofilament light, and glial fibrillary factor), and hypernitrosylation, this study would have been even more significant. Furthermore, a subset of the patients received escitalopram (25%), sertraline (31.3%), fluoxetine (15.6%), and venlafaxine (10.9%). It has been established that administering antidepressants in vitro to peripheral blood mononuclear cells or whole blood (that is stimulated with LPS+PHA in vitro) decreases the production of proinflammatory cytokines and IFN-γ and increases the production of IL-10 (Xia et al., 1996, Maes et al., 1999). As a result, these latter authors postulated that antidepressants possess intrinsic immunoregulatory properties in vitro.

However, it was not possible to observe these in vitro effects in the serum/plasma of patients with depression, likely due to the fact that ongoing trigger factors stimulate the IRS to remain active (Maes et al., 2011). Similarly, the current investigation failed to identify any discernible impacts of the drug state (including antidepressants and other psychotropic medications) on immune profiles. Similarly, the drug states of the patients did not influence the relationships between immune profiles and SDMD. An unresolved inquiry pertains to whether SDMD, characterized by decreased IL-4 and CIRS, serves as a precursor to subsequent MDMD in certain patients via breakdown of immune tolerance and ultimately IRS activation. Further research should investigate whether individuals with SDMD have the potential to develop subsequent MDMD episodes, or if these are in fact two separate categories characterized by distinct immune pathophysiologies.

## Conclusions

SDMD and FE-SDMD are distinguished by the substantial inhibition observed in the alternative M2 macrophage, Th-2, and CIRS profiles. Additionally, SDMD is associated with decreased levels of negative immunoregulatory cytokines IL-4 and IL-10 in the serum, as well as sIL-2R and IL-12p40, which typically provide negative feedback on Th-1-like activities, and the anti-inflammatory growth factor M-CSF. The association between decreased CIRS, IL-4, and sIL-2R levels, and elevated CCL11 and G-CSF levels, accounts for a greater proportion of the variability observed in suicidal behaviors, depression severity, and the SDMD phenotype. Additionally, IL-4 is inversely related to ruminating, a crucial component of the clinical manifestation of SDMD.

Overall, SDSM is associated with compromised immune homeostasis and tolerance, as evidenced by the suppression of the CIRS profile rather than an inflammatory response. Despite the fact that MDMD and SDMD have immunological profiles that differ significantly from one another, both subtypes of depression have an increased IRS/CIRS ratio during their acute stages. This is because the CIRS is either depleted in a relative manner (in the case of MDMD) or in an actual manner (in the case of SDMD). Widespread disruptions in immune, redox, and detoxification mechanisms may result from the suppression of the CIRS profile, which also affects the homeostasis of systems such as microbial defenses, energy metabolism, immune tolerance, and gut-brain axis functions.

As a result, reduced CIRS, IL-4, and IL-10 activities have emerged as novel therapeutic targets for SDMD. In subsequent investigations into depression, it is imperative that a clear distinction be made between both depression subtypes and between the acute and remission phases of both subtypes. Failing to distinguish between these subgroups significantly complicates the interpretation of the results.

## Data Availability

MM will reply to reasonable requests for the dataset used in the current study after it has been fully utilized by all authors.

## Ethical approval and consent to participate

The research project (IRB no.351/63) was approved by the Institutional Review Board of Chulalongkorn University’s institutional ethics board, Bangkok, Thailand, which follows the International Guideline for Human Research protection as required by the Declaration of Helsinki, The Belmont Report, CIOMS Guideline and International Conference on Harmonization in Good Clinical Practice (ICH-GCP). All participants signed the appropriate institutional informed consent forms before data collection.

## Credit author’s contributions

AV and MM carried out the current study’s design. The data was gathered by AV. The statistical evaluation was performed by MM. MM wrote the first draft. All authors contributed to the editing of the work, and they have all given their consent for submission of the completed version.

## Acknowledgments

Not applicable.

