## supplementary file for "Major depression is not an inflammatory disorder: depletion of the compensatory immunoregulatory system is a hallmark of a mild depression phenotype"

### ELECTRONIC SUPPLEMENTARY FILE (ESF)

**Corresponding author:**

China

https://scholar.google.com/citations?user=1wzMZ7UAAAAJ&hl=en

**ESF, Table 1**. Overview of the cytokines, chemokines, and growth factors measured in the current study

| Protein abbreviations | Gene Symbol | > OOR (%) | Protein name / alias |
|---|---|---|---|
| **IFN-α2** | **IFNA2** | 50 | Interferon-α2 |
| **IFN-γ** | **IFNG** | 93 | Interferon-γ |
| **IL-1α** | **IL1A** | 54 | Interleukin-1α |
| **IL-1β** | **IL1B** | 53 | Interleukin-1β |
| **sIL-1RA** | **IL1RN** | 84 | Soluble interleukin-1 receptor antagonist |
| **IL-2** | **IL2** | 50 | Interleukin-2 |
| **IL-2R** | **IL2RA** | 100 | Soluble interleukin-2 receptor |
| **IL-3** | **IL3** | 50 | Interleukin-3 |
| **IL-4** | **IL4** | 100 | Interleukin-4 |
| **IL-5** | **IL5** | 53 | Interleukin-5 |
| **IL-6** | **IL6** | 53 | Interleukin-6 |

| | | | |
|---|---|---|---|
| **IL-7** | **IL7** | 52 | Interleukin-7 |
| **IL-9** | **IL9** | 100 | Interleukin-9 |
| **IL-10** | **IL10** | 52 | Interleukin-10 |
| **IL-12p70** | **IL12RB1** | 52 | Interleukin-12 p70 |
| **IL-12p40** | **IL12RB1** | 50 | Interleukin-12 p40 |
| **IL-13** | **IL13** | 73 | Interleukin-13 |
| **IL-15** | **IL15** | 52 | Interleukin-15 |
| **IL-16** | **IL16** | 100 | Interleukin-16 |
| **IL-17** | **IL17A** | 54 | Interleukin-17 |
| **IL-18** | **IL18** | 97 | Interleukin-18 |
| **TNF-α** | **TNF** | 100 | Tumor necrosis factor-α |
| **TNF-β** | **LTA** | 100 | Tumor necrosis factor-β or lymphotoxin-alpha (LT-α) |
| **TRAIL** | **TNFSF10** | 100 | TNF-related apoptosis-inducing ligand (TRAIL) or tumor necrosis factor ligand superfamily member 10 (TNFSF10) |
| **LIF** | **LIF** | 93 | Leukemia inhibitory factor or IL-6 famiy cutokine |
| **MIF** | **MIF** | 100 | Macrophage migration inhibitory factor-like protein (MIF) or glycosylation-inhibiting factor |

| | | | |
|---|---|---|---|
| **G-CSF** | **CSF3** | 100 | Granulocyte colony stimulating factor (G-CSF) or colony stimulating factor 3 (CSF3) |
| **M-CSF** | **CSF1** | 100 | Macrophage colony-stimulating factor (M-CSF) or colony stimulating factor 1 (CSF1) |
| **GM-CSF** | **CSF2** | 66 | Granulocyte-macrophage colony-stimulating factor (GM-CSF) or colony-stimulating factor 2 (CSF2) |
| **CCL2 or MCP1** | **CCL2** | 100 | C-C motif chemokine ligand 2 (CCL2) or monocyte chemoattractant protein 1 (MCP1) |
| **CCL3 or MIP-1α** | **CCL3** | 94 | C-C motif Chemokine ligand 3 (CCL3) or macrophage inflammatory protein 1-alpha (MIP-1α) |
| **CCL4 or MIP-1β** | **CCL4** | 100 | C-C motif chemokine ligand 4 (CCL4) or macrophage inflammatory protein 1β (MIP-1β) or lymphocyte activation gene 1 protein |
| **CCL5 or RANTES** | **CCL5** | 100 | C-C motif chemokine ligand 5 (CCL5) or regulated upon activation, normally T-expressed, and presumably Secreted (RANTES) |
| **CCL7 or MCP3** | **CCL7** | 53 | C-C motif chemokine ligand 7 (CCL7) or monocyte-chemotactic protein 3 (MCP3). |
| **CCL11 or Eotaxin** | **CCL11** | 100 | C-C motif chemokine ligand 11 (CCL11) or eosinophil chemotactic protein |
| **CCL27 or CTACK** | **CCL27** | 100 | C-C motif chemokine ligand 27 (CCL27) or cutaneous T-cell attracting chemokine (CTACK) |
| **CXCL1 or GRO-α** | **CXCL1** | 100 | C-X-C motif chemokine 1 (CXCL1) or growth-regulated alpha protein (GRO) |
| **CXCL8 or IL-8** | **CXCL8** | 100 | C-X-C motif chemokine ligand 8 (CXCL8) or interleukin-8 (IL-8) |
| **CXCL9 or MIG** | **CXCL9** | 100 | C-X-C motif chemokine ligand 9 (CXCL9) or monokine induced by gamma interferon (MIG) |
| **CXCL10 or IP10** | **CXCL10** | 100 | C-X-C motif chemokine ligand 10 (CXCL10) or Interferon gamma-induced protein 10 (IP10) |
| **CXCL12 or SDF-1α** | **CXCL12** | 100 | C-X-C motif chemokine 12 (CXCL12) or stromal cell-derived factor 1 (SDF-1α) |

| | | | |
|---|---|---|---|
| **FGF** | **FGF2** | 37 | Fibroblast growth factor 2 (FGF) or basic fibroblast growth factor |
| **HGF or SF** | **HGF** | 100 | Hepatocyte growth factor (HGF) or scatter factor (SF) |
| **BNGF** | **NGF** | 62 | β-nerve growth factor (NGF) |
| **PDGF** | **PDGFA** | 100 | Platelet derived growth factor (PDGF) |
| **SCF** | **KITLG** | 100 | Stem cell factor (SCF) or Kit ligand (KITLG) |
| **SDF-1** | **SDF1** | 100 | Stromal cell-derived factor |
| **SCGF-β or CLEC11A** | **CLEC11A** | 86 | Stem cell growth factor (SCGF) or C-type lectin domain family 11 member A (CLEC11A) |
| **VEGF** | **VEGFA** | 38 | Vascular endothelial growth factor (VEGF) |

Adapted from: *Maes M, Rachayon M, Jirakran K, Sodsai P, Klinchanhom S, Gałecki P, Sughondhabirom A, Basta-Kaim A. The Immune Profile of Major Dysmood Disorder: Proof of Concept and Mechanism Using the Precision Nomothetic Psychiatry Approach. Cells. 2022 Mar 31;11(7):1183. doi: 10.3390/cells11071183. PMID: 35406747; PMCID: PMC8997660.*

*Kalayasiri R, Dadwat K, Supaksorn T, ,Sirivichayakul S, Maes M. Methamphetamine (MA) use, MA dependence, and MA-induced psychosis are associated with increasing aberrations in the compensatory immunoregulatory system and interleukin-1α and CCL5 levels. medRxiv 2023.03.26.23287766; doi: https://doi.org/10.1101/2023.03.26.23287766*

**ESF, Table 2**. Description of the immune profiles used in this study

| Immune Profile | Members |
|---|---|
| **M1 macrophage** | IL-1β, IL-6, TNF-α, IL-12p70, IL-15, CCL2, CCL5, CXCL1, CXCL8, CXCL9, CXCL10 |
| **M2 macrophages** | IL-10, IL-4, IL-13, VEGF, PDGF, sIL-1RA |
| **z M1 – z M2** | zM1 – zM2 |
| **T helper (Th)-1** | IL-2, sIL-2R, IFN-a, IFN-γ, IL-12p70, IL-16, TNF-α, TNF-β |
| **Th-2** | IL-4, IL-5, IL-9, IL-13, IL-10, IL-6 |
| **z Th- z Th-2** | zTh-1 – zTh-2 |
| **IRS** | IL-1α, IL-1β, IL-6, TNF-α, IL-12p70, IL-15, IL-16, IL-17, IL-18, CCL2, CCL3, CCL4, CCL5, CCL7, CCL11, CXCL1, CXCL8, CXCL9, CXCL10, IL-2, IFN-α, IFN-γ, TNF-α, TNF-β, TRAIL, GM-CSF, M-CSF, G-CSF, SCGF |
| **CIRS** | IL-4, IL-10, sIL-1RA, sIL-2R |
| **z IRS – z CIRS** | zIRS – zCIRS |

IRS: immune-inflammatory response system; CIRS: compensatory immunoregulatory system

Adapted from:

*Maes M, Rachayon M, Jirakran K, Sodsai P, Klinchanhom S, Gałecki P, Sughondhabirom A, Basta-Kaim A. The Immune Profile of Major Dysmood Disorder: Proof of Concept and Mechanism Using the Precision Nomothetic Psychiatry Approach. Cells. 2022 Mar 31;11(7):1183. doi: 10.3390/cells11071183. PMID: 35406747; PMCID: PMC8997660.*

*Kalayasiri R, Dadwat K, Supaksorn T, ,Sirivichayakul S, Maes M. Methamphetamine (MA) use, MA dependence, and MA-induced psychosis are associated with increasing aberrations in the compensatory immunoregulatory system and interleukin-1α and CCL5 levels. medRxiv 2023.03.26.23287766; doi: https://doi.org/10.1101/2023.03.26.23287766*

**ESF, Table 3**. Differences in immune profiles between students with a first-episode simple dysmood disorder (FE-SDMD) and healthy students (HC).

**Estimates**

| Dependent Variable | FE-SDMD versus HC | Mean | Std. Error | 95% Confidence Interval | |
|---|---|---|---|---|---|
| | | | | Lower Bound | Upper Bound |
| M1 | 0 | .101[a] | .154 | -.205 | .406 |
| | 1 | .060[a] | .148 | -.235 | .356 |
| M2 | 0 | .202[a] | .157 | -.111 | .514 |
| | 1 | -.100[a] | .152 | -.402 | .202 |
| zM1-zM2 | 0 | -.101[a] | .159 | -.417 | .215 |
| | 1 | .161[a] | .153 | -.144 | .466 |
| Th-1 | 0 | .246[a] | .155 | -.062 | .554 |
| | 1 | -.042[a] | .150 | -.340 | .255 |
| Th-2 | 0 | .329[a] | .145 | .040 | .619 |
| | 1 | -.130[a] | .140 | -.409 | .149 |
| zTh1-zTh2 | 0 | -.083[a] | .149 | -.380 | .214 |
| | 1 | .088[a] | .144 | -.199 | .374 |
| IRS | 0 | .145[a] | .153 | -.158 | .449 |
| | 1 | .020[a] | .147 | -.273 | .313 |
| CIRS | 0 | .405[a] | .146 | .114 | .696 |
| | 1 | -.209[a] | .141 | -.490 | .072 |
| zIRS-zCIRS | 0 | -.260[a] | .127 | -.513 | -.006 |
| | 1 | .229[a] | .123 | -.016 | .474 |

a. Covariates appearing in the model are evaluated at the following values: Age = 22.6873, Sex = .14, BMI = 22.0432, Current use_smoking = .07, Regular drinking_current (1 mo.) = .13.

#### Univariate Tests

| Dependent Variable | | Sum of Squares | df | Mean Square | F | Sig. |
|---|---|---|---|---|---|---|
| M1 | Contrast | .033 | 1 | .033 | .034 | .855 |
| | Error | 82.552 | 84 | .983 | | |
| M2 | Contrast | 1.865 | 1 | 1.865 | 1.814 | .182 |
| | Error | 86.376 | 84 | 1.028 | | |
| zM1-zM2 | Contrast | 1.402 | 1 | 1.402 | 1.338 | .251 |
| | Error | 88.045 | 84 | 1.048 | | |
| Th-1 | Contrast | 1.700 | 1 | 1.700 | 1.700 | .196 |
| | Error | 83.999 | 84 | 1.000 | | |
| Th-2 | Contrast | 4.313 | 1 | 4.313 | 4.899 | .030 |
| | Error | 73.944 | 84 | .880 | | |
| zTh1-zTh2 | Contrast | .597 | 1 | .597 | .643 | .425 |
| | Error | 77.946 | 84 | .928 | | |
| IRS | Contrast | .320 | 1 | .320 | .330 | .567 |
| | Error | 81.435 | 84 | .969 | | |
| CIRS | Contrast | 7.711 | 1 | 7.711 | 8.651 | .004 |
| | Error | 74.872 | 84 | .891 | | |
| zIRS-zCIRS | Contrast | 4.889 | 1 | 4.889 | 7.234 | .009 |
| | Error | 56.774 | 84 | .676 | | |

The F tests the effect of FE-SDMD vs HC.

**ESF, Table 4.** Differences in cytokines and growth factors between students with simple dysmood disorder (SDMD) and healthy students (HC).

**Estimates**

| Dependent Variable | hc_FIRSTEPISODE | Mean | Std. Error | 95% Confidence Interval | |
|---|---|---|---|---|---|
| | | | | Lower Bound | Upper Bound |
| sIL2R | 0 | .261[a] | .151 | -.040 | .562 |
| | 1 | -.159[a] | .146 | -.450 | .132 |
| IL-4 | 0 | .344[a] | .152 | .043 | .646 |
| | 1 | -.201[a] | .146 | -.492 | .091 |
| IL-10 | 0 | .321[a] | .161 | .001 | .641 |
| | 1 | -.193[a] | .155 | -.502 | .116 |
| IL12p40 | 0 | .315[a] | .148 | .020 | .611 |
| | 1 | -.158[a] | .143 | -.443 | .127 |
| M-CSF | 0 | .305[a] | .158 | -.010 | .620 |
| | 1 | -.135[a] | .153 | -.439 | .169 |

a. Covariates appearing in the model are evaluated at the following values: Age = 22.6873, Sex = .14, BMI = 22.0432, Current use_smoking = .07, Regular drinking_current (1 mo.) = .13.

**Univariate Tests**

| Dependent Variable | | Sum of Squares | df | Mean Square | F | Sig. |
|---|---|---|---|---|---|---|
| sIL-2R | Contrast | 3.601 | 1 | 3.601 | 3.774 | .055 |
| | Error | 80.164 | 84 | .954 | | |
| IL-4 | Contrast | 6.068 | 1 | 6.068 | 6.341 | .014 |
| | Error | 80.384 | 84 | .957 | | |
| IL-10 | Contrast | 5.394 | 1 | 5.394 | 5.011 | .028 |
| | Error | 90.411 | 84 | 1.076 | | |
| IL-12p40 | Contrast | 4.584 | 1 | 4.584 | 5.004 | .028 |
| | Error | 76.948 | 84 | .916 | | |
| M-CSF | Contrast | 3.962 | 1 | 3.962 | 3.798 | .055 |
| | Error | 87.632 | 84 | 1.043 | | |

The F tests the effect of FE-SDMD vs HC.
